# A phase I/II trial to treat massive Africanized honeybee (*Apis mellifera*) stings using the new apilic antivenom

**DOI:** 10.1101/2020.12.26.20248062

**Authors:** Alexandre Naime Barbosa, Rui Seabra Ferreira, Francilene Capel Tavares de Carvalho, Fabiana Schuelter-Trevisol, Mônica Bannwart Mendes, Bruna Cavecci Mendonça, José Nixon Batista, Daisson José Trevisol, Leslie Boyer, Jean-Philippe Chippaux, Natália Bronzatto Medolago, Claudia Vilalva Cassaro, Márcia Tonin Rigotto Carneiro, Ana Paola Piloto de Oliveira, Daniel Carvalho Pimenta, Luís Eduardo Ribeiro da Cunha, Lucilene Delazari dos Santos, Benedito Barraviera

**Affiliations:** Department of Infectology, Dermatology, Imaging Diagnosis and Radiotherapy, Botucatu Medical School (FMB), São Paulo State University (UNESP – Univ Estadual Paulista), Botucatu, SP, Brazil; Graduate Program in Tropical Diseases, Botucatu Medical School (FMB), São Paulo State University (UNESP – Univ Estadual Paulista), Botucatu, SP, Brazil; Center for the Study of Venoms and Venomous Animals (CEVAP), São Paulo State University (UNESP – Univ Estadual Paulista), Botucatu, SP, Brazil; Clinical Research Center at Hospital Nossa Senhora da Conceição, Tubarão, SC, Brazil; Graduate Program in Health Sciences, University of Southern Santa Catarina at Tubarão, SC, Brazil; VIPER Institute, University of Arizona College of Medicine, Tucson, AZ USA; MERIT, IRD, Université Paris 5, Sorbonne Paris Cité, Paris, 75006, France; CRT, Institut Pasteur, Paris 75015, France; Clinical Research Unit (UPECLIN), Botucatu Medical School, São Paulo State University (UNESP – Univ Estadual Paulista), Botucatu, SP Brazil; Butantan Institute, São Paulo, SP, Brazil; Vital Brazil Institute, Niterói, RJ, Brazil; Graduate Program in Clinical Research, Center for the Study of Venoms and Venomous Animals (CEVAP), São Paulo State University (UNESP – Univ Estadual Paulista), Botucatu, SP, Brazil

**Keywords:** Antivenom, *Apis mellifera* Africanized, Clinical trial, Safety assessment, ELISA - Enzyme-linked immunosorbent assay

## Abstract

Safety, optimal minimum dose, and, preliminary effectiveness of a new generation Africanized honeybees (*Apis mellifera*) antivenom (AAV) were evaluated. A phase I/II, multicenter, non-randomized, single-arm clinical trial involving 20 participants showing multiple stings were studied. Participants have received either 2 to 10 vials of AAV based on the stings number together with a predefined adjuvant, symptomatic, and complementary treatment schedule. The primary safety endpoint was the presence of early adverse reactions within the first 24 hours after treatment. Preliminary efficacy through clinical evolution, including laboratory tests, was assessed at baseline and over the following four weeks. ELISA assays and mass spectrometry estimated the venom pharmacokinetics before, during, and after treatment. Twenty adult participants, 13 (65%) males, and 7 (35%) females, with a median age of 44 years and a mean body surface of 1.92 m^2^ (median = 1.93 m^2^) were recruited. The median number of stings was 52.5 ranging from 7 to more than 2,000. Envenoming severity was classified as 80% mild, 15% moderate, and 5% severe. According to the protocol, 16 (80%) participants received two AAV vials, 3 (15%) six vials, and one (5%) 10 vials. There was no discontinuation of the treatment due to acute adverse events and there were no late adverse reactions. Two patients showed mild adverse events with only transient itchy skin and erythroderma. All participants completed the infusion within two hours and there was no loss of follow-up after discharge. ELISA assays showed venom concentrations varying between 0.25 ng/mL and 1.479 ng/mL prior to treatment. Venom levels decreased in all cases during the hospitalization period. Surprisingly, in nine cases (45%), despite clinical recovery and without symptoms, the venom levels increased again during outpatient care 10 days after discharge. Mass spectrometry showed melittin in eight participants 30 days after the treatment. Considering the promising safety results of the investigational product for the treatment of massive Africanized honeybee attacks, added to efficacy in clinical improvement and immediate decrease in blood venom level, the AAV has shown to be safe for human use.

**Trial registration:** Universal Trial Number (UTN): U1111-1160-7011, Register Number: RBR-3fthf8 (http://www.ensaiosclinicos.gov.br/rg/RBR-3fthf8/).

## 1. Introduction

In 1956, African honeybees of the species *Apis mellifera scutellata* were introduced from Tanganyika and South Africa to Brazil because they were more productive and more resistant to pests (1). They accidentally escaped and crossed with existing European bees of the species *Apis mellifera mellifera*, originating since then an Africanized hybrid. This present marked defensive, great swarming capacity, and easy adaptation to different climates and environments. In addition, it made it possible to expand throughout Brazil and several other countries in the Western Hemisphere, including nowadays the United States (2, 3, 4, 5). These honeybees, when molested, massive attack the target and, consequently, the number of accidents with human beings and animals since then has increased. (4, 6, 7). These honeybees, when disturbed, massive attack their target resulting in an increase, since 1956, the number of accidents in humans and animals.

Bee venom is composed of complex mixtures of biogenic amines, proteins, enzymes and peptides. Among the peptides of low allergenic importance, but of intense pharmacological action, there are melittin, which constitutes 50 to 60% of the gross weight, phospholipase A_2_ comprising about 11 to 12%, apamine (3%), hyaluronidase (1%-2%), several low molecular weight peptides (1%), water, and mineral salts. Note that melittin acts synergistically with phospholipase A_2_, acting as a diffusion factor for the venom, and therefore, comprising its two most toxic components (8, 9,10).

In the scope of venomous animals, envenoming by venomous snakes is a serious public health problem for tropical countries, being one of the neglected diseases classified by the World Health Organization (WHO) (11, 12). This becomes more serious when the envenomation is caused by the fairly large amounts of venom injected, such as biting by *Bothrops jararaca* adult. Similarly, can also be the case for *Apis mellifera* massive attacks when thousands of bees will inject up to half a gram of venom. The expansion of Africanized honeybees (AHB) throughout the Americas (2, 4, 5, 13) and consequently the severity of their accidents also forced health authorities to classify such accidents as an object of health surveillance as in Brazil (**Figure 1**) (14).

**Figure 1:**
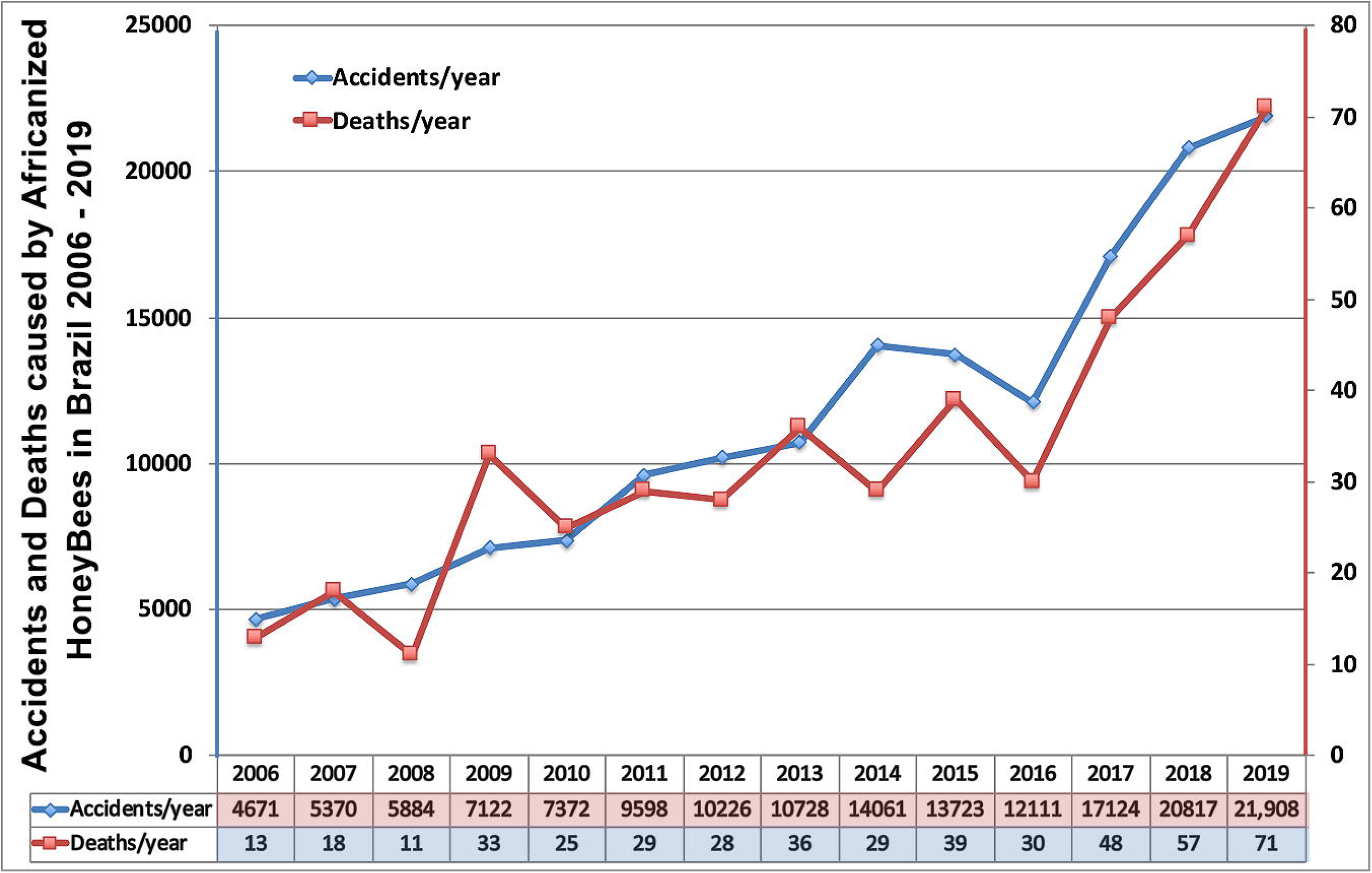
Annual distribution of the number of accidents and deaths by AHB in Brazil between 2006 and 2019 (14).

These accidents show clinical manifestations that depend mainly on the number of stings but also on the individual’s sensitivity. The most frequent accident is one where the individual not sensitized to venom is affected by a few stings. In these cases, the clinical presentation is limited only to the local inflammatory reaction, which is manifested by erythematous papules, pain, and heat. In most cases, the situation is resolved without medical participation. Another clinical form is that in which the individual sensitized to one or more components of the venom, exhibits an immediate type I hypersensitivity reaction, as defined by Coombs and Gell (15). This is a serious event that can be triggered by only one sting, requiring immediate medical intervention. Generally, the clinical manifestations are glottal edema, or angioedema, and bronchospasm associated with anaphylactic shock (4, 6, 7).

The third form of presentation is that caused by multiple stings occurring when the individual is attacked by a swarm. Here, a large amount of venom is inoculated, usually caused by hundreds or thousands of honeybees (4, 6, 7, 16-18). These participants present generalized pain, intense itching, and agitation, which may progress to numbness, associated with severe acute respiratory and kidney failures. Those who evolve to death present on pathological examination with acute tubular necrosis as well as heme and/or myoglobin cylinders inside the renal tubules or glomeruli. Skeletal muscles exhibit intense proteolysis with the release of myoglobin and creatine phosphokinase (CPK) into the blood. The heart may be affected and, in this case, a subendocardial lesion with the presence of infarction is observed. The liver may show signs of hydropic degeneration due to severe envenoming (4, 7, 16-18).

Laboratory tests change rapidly and the blood count may show leukocytosis with neutrophilia and a staggered leftward shift; examination of type I urine reveals proteinuria, glycosuria, and heme pigment. Serum levels of urea and creatinine may increase due to kidney damage. Levels of CPK and aspartate aminotransferase (AST) are usually elevated due to severe rhabdomyolysis. The levels of alanine aminotransferase (ALT) may rise over time, indicating liver failure. Finally, acute-phase proteins, including C-reactive protein (CRP) and fibrinogen, are altered, reflecting a severe systemic inflammatory response syndrome (4, 6, 7, 18).

Until recently, the treatment of both small and large numbers of stings was symptomatic and relied on antihistamines, corticosteroids, and even epinephrine for anaphylactic shock. The search for a specific treatment based on heterologous antivenom was a challenge for many researchers (19, 20). In 2000, Ferreira Jr. et al. (4, 21, 22) initiated the development of a new antivenom constituted only by antibodies against the two main toxins, namely melittin and phospholipase A_2_. In 2017, Barbosa et al. (23) published a clinical protocol for the treatment of participants with multiple stings, which was applied in this clinical study.

This study was performed a) to assess the safety, b) to establish the optimal minimum dose, and, c) to evaluate the preliminary effectiveness of the novel Apilic Antivenom (AAV).

## 2. Patients and Methods

### 2.1 Ethics statement

The clinical protocol has been previously approved by the Brazilian National Commission on Ethics in Research (CONEP, Certificate of Presentation of Ethical Appreciation No. 19006813.4.1001.5411, v7, approved in 06/07/2016) and the Brazilian National Health Surveillance Agency (ANVISA) whose Consent Record of Apis Study was approved on 02/05 2016 by No. 0907532142, Proc. No. 25361611582201493. This trial RBR-3FTHF8 was registered in 2015 in the Brazilian Clinical Trials Registry (ReBEC) at http://www.ensaiosclinicos.gov.br/. The first participant was included in 08/22/2016, the Universal Trial Number (UTN) is U1111-1160- 7011, the Register Number is RBR-3fthf8 and the public access URL is available at http://www.ensaiosclinicos.gov.br/rg/RBR-3fthf8/ (24) The clinical trial protocol was published by Barbosa *et al*. in 2017 (23) and is available at https://doi.org/10.1186/s40409-017-0106-y.

### 2.2 Study Design

The phase I/II, multicenter, non-randomized, single-arm clinical trial involving 20 multiple- sting participants by AHB treated with the new Apilic Antivenom “*batch 155804 R*” (**Figure 2**). Two clinical research units belonging to the Brazilian National Clinical Research Network (RNPC) were requested for this study, namely: UNESP – Botucatu, Brazil and HNSC – Tubarao, Brazil. All participants who consented to participate signed the Free and Informed Consent Form (FICF).

**Figure 2:**
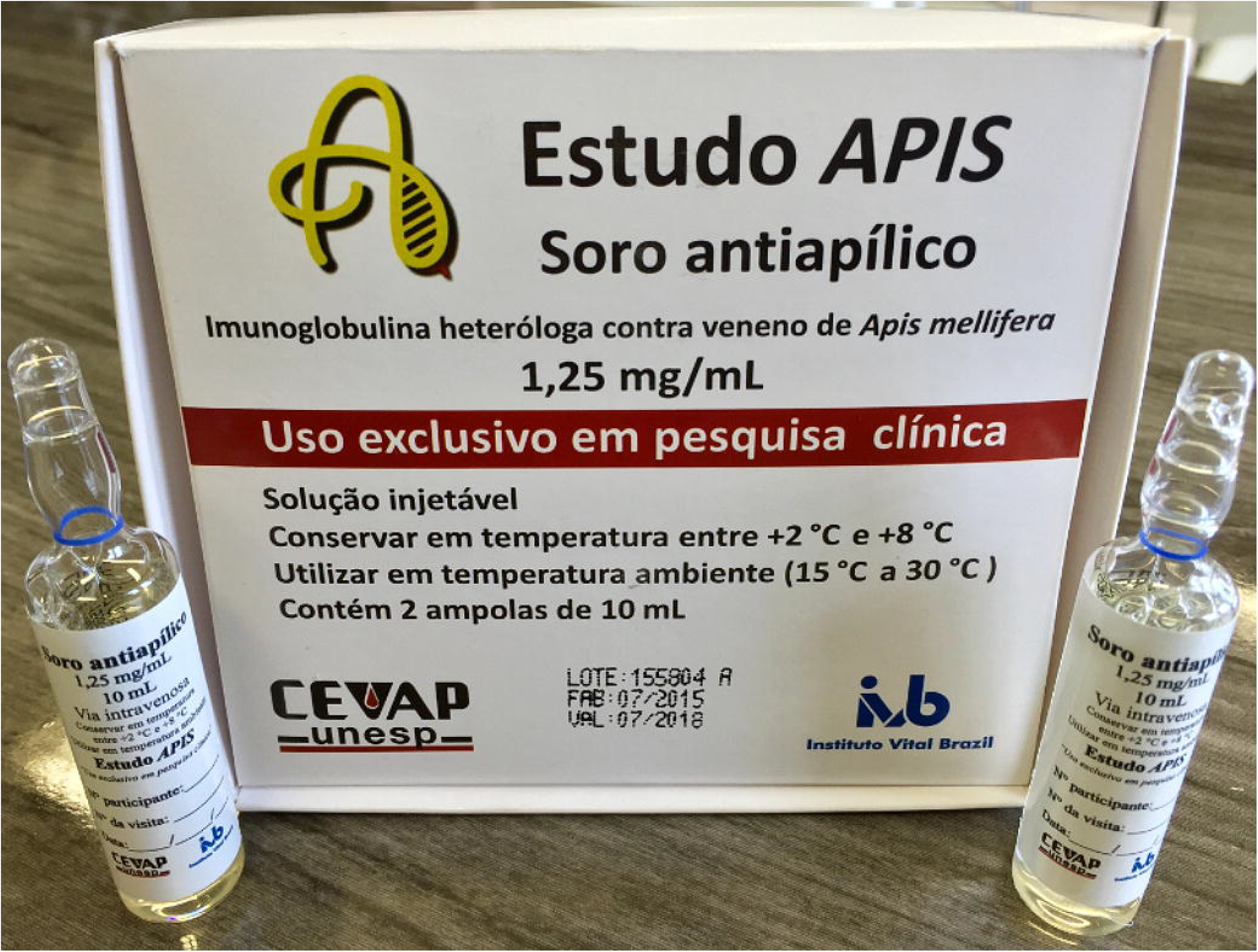
New Apilic Antivenom (AAV)

### 2.3 Outcomes

The primary endpoints were to assess the safety profile of AAV in participants exposed to multiple stings from Africanized honeybees based on adverse event occurrences and estimate the proportion of participants showing improvement over to the initial clinical state by monitoring symptoms and laboratory tests.

The secondary endpoint was to evaluate the correlation between the number of stings and the severity of the initial clinical picture by the AHB score based in APACHE II and adapted for this envenoming (25).

In addition, an exploratory endpoint was included in the study to estimate the pharmacokinetic profile of AHB venom by ELISA tests on blood samples collected at different moments (before, and at 2, 6, 12, and 24 h after AAV administration). The aim was to assess the acute-phase-reaction profile of AVV as determined by the variations of the C-reactive protein (CRP) and fibrinogen levels.

### 2.4 Inclusion, exclusion, and discontinuation criteria

All participants who presented after AHB stings were screened for eligibility by the clinical staff present according to **Table 1**. People over 18 years of age were eligible for AVV use according to the number of stings, as described below. Each participant or a relative signed the FICF.

**Table 1.** Inclusion, exclusion, and discontinuation criteria.

| Inclusion | Exclusion | Discontinuation |
| --- | --- | --- |
| Participants over 18 years of age of both sexes | Participants who had a previous adverse reaction to heterologous serum produced in horses. | Developing anaphylactic shock resistant to the management protocol for reactions of acute hypersensitivities. |
| Participants admitted to the hospital after an accident with AHB | Participants who are pregnant or nursing | Withdrawing from the terms of free and informed consent (TFIC). |
| Participants or a responsible relative who signs informed consent to receive the antivenom. | - | - |

### 2.5 Antivenom doses and adjuvant treatments

Before treatment, the following procedures were carried out: height was measured in centimeters (cm), body weight in kilograms (kg) body mass index in Kg/m^2^; and body surface area in square meters (m^2^), as described by Madden and Smith (26). Then, all participants received AAV in a single administration, diluted in a solution of 250 mL of 0.9% sodium chloride, by intravenous route over two hours, according to the protocol and number of stings described below:

- Up to 5 stings: specific treatment with Apilic Antivenom was not indicated. Here, only adjuvant, symptomatic and complementary treatments were applied;
- Between 5 and 200 stings: two vials (20 ml) of Apilic Antivenom;
- Between 201 and 600 stings: six vials (60 ml) of Apilic Antivenom;
- Above 600 stings: ten vials (100 ml) of Apilic Antivenom.

The adjuvant, symptomatic and complementary treatments were described and published in detail by Barbosa et al. (23) at https://doi.org/10.1186/s40409-017-0106-y.

### 2.6 Criteria for severity and clinical outcome of the participants

An AHB score was assigned, based on APACHE II, which is a validated system for classifying severity of the disease (25). The proposed AHB score varied between 1 and 15 according to the severity criteria that included eight clinical and seven strategic laboratory alterations, and was assessed at the time of the first examination and based on the medical literature (4, 7, 16-18). The same criteria, except for those that did not change during the treatment (age over 60, body mass index over 30 kg/m^2^, time elapsed between the accident and medical care, and number of stings) were evaluated and monitored during the course of the envenoming to verify the clinical outcome and normalization of laboratory tests upon discharge from the hospital and during follow-up visits, 10, 20, and 30 days afterward.

#### 2.6.1 AHB Score proposed for clinical and laboratory assessments

– Age over 60 years – score 1
– Body mass index over 30 kg/m^2^ – score 1
– Time elapsed between the accident and medical care (over 24 hours) – score 1
– Number of stings
  - 5-200 stings - score1
  - 201-600 stings - score 2
  - More than 600 stings - score 3
– Hemodynamic disorders (tachycardia, arterial hypotension, shock) score 1
– Respiratory disorders (bradypnea, bronchospasm, wheezing and/or dyspnea) – score 1
– Neurological disorders (mental confusion and/or intense headache) – score 1
– Acute kidney failure (anuria and/or oliguria) – score 1
– Rise in levels de creatine phosphokinase (CPK) – score 1
– Increase in levels of alanine amino transferase (ALT) – score 1
– Rise in levels of creatinine – score 1
– Elevation in levels of C-reactive protein (CRP) – score 1
– Increase in fibrinogen levels – score 1
– Increase in leukocyte levels – score 1
– Diminution in platelet count – score 1

#### 2.6.2 Subsidiary and strategic laboratory tests

Laboratory tests (CPK, ALT, Creatinine, CRP, Fibrinogen, Leukocytes and, Platelets) were used to assess the safety parameters before treatment (hospital admission), on discharge day and at follow-up (10, 20 and 30 days after hospital discharge). All the procedures and protocol for laboratory tests are presented in **Additional File 1** (27, 28).

#### 2.6.3 Classification of acute adverse reactions to antivenom

Adverse reactions to AAV were predefined as mild, moderate, or severe based on an international classification of the anaphylaxis reactions described in **Table 2** (29, 30).

**Table 2.**
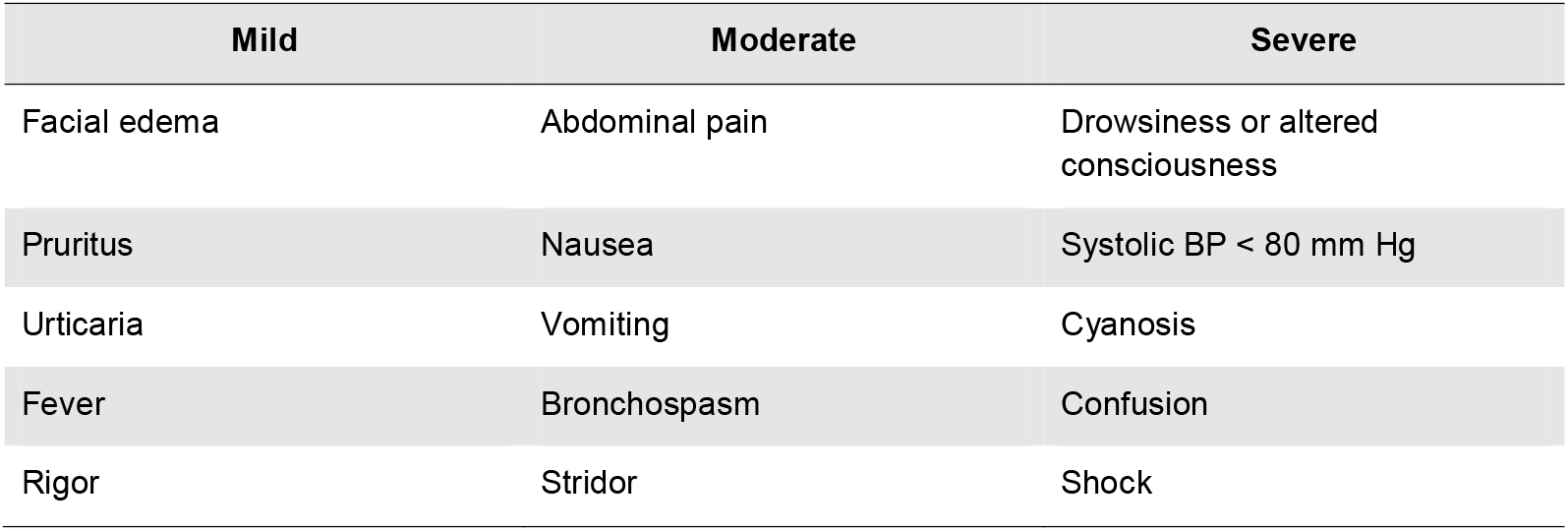
Classification of acute adverse reactions to antivenom (29, 30).

| Mild | Moderate | Severe |
| --- | --- | --- |
| Facial edema | Abdominal pain | Drowsiness or altered consciousness |
| Pruritus | Nausea | Systolic BP < 80 mm Hg |
| Urticaria | Vomiting | Cyanosis |
| Fever | Bronchospasm | Confusion |
| Rigor | Stridor | Shock |

### 2.7. ELISA assays to estimate venom pharmacokinetics

F(ab’)_2_ type immunoglobulin fractions prepared from the blood of hyperimmunized horses against the venom melittin and PLA_2_ molecules of the AHB of the species *Apis mellifera* [Anti- melittin F(ab’)_2_ and Anti-PLA_2_ F(ab’)_2_] were obtained from the AAV using a single affinity chromatographic step as described by Chávez-Olórtegui et al. modified (31). Thirty-two milligrams of crude venom from *A. mellifera* Africanized honeybees were immobilized with 3 grams of CNBr-Sepharose resin and prepared according to the manufacturer’s instructions (Cytiva) for the aforementioned affinity chromatographic assay. Anti-melittin F(ab’)_2_ and Anti-PLA_2_ F(ab’)_2_ were conjugated to peroxidase (HRP - Sigma) according to the method of Nakane and Kawoi (32). The conjugate was titrated as described by Chávez-Olórtegui et al. (31) modified with the conjugate being diluted at the proportions 1:20, 1:2,000, 1:5,000, and 1:10,000, and its viability evaluated for 30 min.

Then, an ELISA assay was performed to quantify the melittin and PLA_2_ fractions in the blood of participants, according to the methodology of Bucaretchi et al. (33). For this, plates containing 96 wells were sensitized with 100 μL of a mixture of Anti-melittin F(ab’)_2_ and Anti- PLA_2_ F(ab’)_2_ at a concentration of 20 μg/mL. The conjugate was used at 1:2000 dilution, both in the calibration curve containing crude venom from *Apis mellifera* and in the blood samples of the participants. The calculations were performed using software Excel in which linear regression analysis and dose-response curve were used. The results were expressed as ng/mL.

### 2.8 Mass spectrometry analyses

Participants’ serum (50 µL) was added to 5% DMSO, 0.1% acetic acid (50 µL) and vortexed for 30 min. After that, the solution was centrifuged for 3 minutes at 3000 g. The supernatant was collected for processing. Purified melittin from crude *Apis mellifera* venom (8) was employed as a standard for method development.

Samples were analyzed by liquid chromatography-mass spectrometry in an ESI-IT-TOF instrument coupled to an UFLC 20A Prominence (Shimadzu, Kyoto, Japan). Samples of 15 μL were injected into a C18 column (Kinetex C18 2.6 µm 100 Å, 100 × 4.6 mm) and analyzed by a binary gradient, employing as solvents: (A) water: DMSO: acetic acid (949: 50: 1) and (B) ACN: DMSO: water: acetic acid (850: 50: 99: 1). Melittin optimal detection conditions were achieved by an elution gradient of 25%-50% B for 20 min at a constant flow of 0.7 mL.min^-1^, after initial isocratic elution for 5 min. Eluates were monitored by a Shimadzu SPD-M20A PDA detector before being injected into the mass spectrometer.

The interface was maintained at 4.5 kV and 275°C. The detector voltage was 1.95 kV and the fragmentation were induced by an argon collision, with 55 ‘energy’ parameters. MS spectra were acquired in positive mode in the m/z range of 700-730 and MS/MS spectra were collected in the range of 50 to 1400 m/z according to previous optimization with purified melittin. The m/z ion 712.15 (M+4H^+^) was selected for fragmentation and [y_13_]^2+^ ion (811.95, the tallest peak) was monitored at the MS^2^ (Supplemental material at **Additional File 2)**.

### 2.9 Statistical analysis

The statistical analysis, as well as the choice of tests for comparison among the research participants were performed with respect to the presuppositions determined by the results, characteristics and course of the variables in the study. The binomial variables were compared by chi-square and Fisher’s exact test. The numerical variables will be compared by the Student’s *t*- test or U test of Mann-Whitney. Statistical analyses of the pharmacokinetic assay were performed via the software GraphPad Prism version 8.3.0, considering differences statistically significant when p<0.05. The results obtained were compared using the ANOVA test for repeated measures, followed by Tukey’s test. The data were represented as mean ± standard error of the mean (34, 35).

## 3 RESULTS

### 3.1 Description of participants

Twenty participants were included, 13 males (65%) and 7 (35%) females, with ages varying between 22 and 77 years, with a median of 44, 19 whites and one brown. The number of stings varied from 7 to more than 2000. The number of vials administered, following the clinical protocol, were: mild cases (2 vials) 16 participants, moderate cases (6 vials) 3 participants, and one participant with a severe case (10 vials). The time elapsed between the accident and the clinical care varied as follows: less than 24 hours = 5 cases, one day = 5 cases, between 2 and 6 days = 8 cases. **Table 3** deals the data.

**Table 3.** Description of participants: study protocol number, age, sex, professional occupation, estimated number of stings, number of vials administered, and the time elapsed between the accident and medical care in days.

| Age | Sex | Occupation | Estimated number of stings | Number of antivenom vials | Time between accident and clinical care |
| --- | --- | --- | --- | --- | --- |
| >30 | F | B | 400 | 6 | 3 |
| >30 | M | B | 40 | 2 | 10 |
| >30 | M | A | 10 | 2 | 0 |
| >20 | M | A | 16 | 2 | 0 |
| >30 | F | A | 10 | 2 | 2 |
| >50 | F | A | 150 | 2 | 1 |
| >50 | M | A | 500 | 6 | 19 |
| >40 | M | A | 55 | 2 | 2 |
| >40 | M | A | 165 | 2 | 2 |
| >30 | F | A | 10 | 2 | 4 |
| >50 | F | B | 30 | 2 | 4 |
| >40 | F | B | 50 | 2 | 4 |
| >30 | M | A | 500 | 6 | 1 |
| >60 | M | B | 100 | 2 | 1 |
| >30 | M | A | 180 | 2 | 1 |
| >40 | M | A | 2000 | 10 | 6 |
| >60 | M | A | 20 | 2 | 0 |
| >70 | M | A | 150 | 2 | 0 |
| >20 | M | A | 7 | 2 | 1 |
| >60 | F | A | 50 | 2 | 0 |

M = male, F = female; Occupation: A – occupation not related to agricultural or wilderness activities; B – occupation related to agricultural or wilderness activities; Colors: White – mild cases; Yellow – moderate cases; Orange – severe case.

**Table 4** details the following data: height varied between 151 and 190 centimeters; body mass index (BMI) ranged from 19.3 to 32.8 kg/m^2^ (mean = 26.4; median = 26.1). Finally, the body surface area (BSA) measured in m^2^, varied from 1.54 to 2.30 (mean = 1.92 m^2^; median = 1.93 m^2^).

**Table 4.** Description of participants: study protocol number, height in centimeters (cm), weight in kilograms (Kg), body mass index in Kg/m^2^, body surface area in m^2^ and estimated number of stings.

| Height in centimeters (cm) | Weight in kilograms (Kg) | Body mass index | Body surface area | Number of stings |
| --- | --- | --- | --- | --- |
| 176 | 65 | 21.0 | 1.78 | 400 |
| 175 | 80 | 26.1 | 1.97 | 40 |
| 178 | 94 | 29.7 | 2.16 | 10 |
| 190 | 100 | 27.7 | 2.30 | 16 |
| 157 | 62,8 | 25.5 | 1.66 | 10 |
| 165 | 81 | 29.8 | 1.93 | 150 |
| 175 | 77 | 25.1 | 1.93 | 500 |
| 179 | 77 | 24.0 | 1.96 | 55 |
| 165 | 52,5 | 19.3 | 1.54 | 165 |
| 168 | 70 | 24.8 | 1.81 | 10 |
| 161 | 67 | 25.8 | 1.73 | 30 |
| 175 | 88 | 28.7 | 2.07 | 50 |
| 188 | 98,1 | 27.8 | 2.26 | 500 |
| 167 | 74 | 26.5 | 1.85 | 100 |
| 182 | 78 | 23.5 | 1.99 | 180 |
| 173 | 89 | 29.7 | 2.07 | 2000 |
| 198 | - | - | - | 20 |
| 168 | 70 | 24.8 | 1.81 | 150 |
| 167 | 91,5 | 32.8 | 2.05 | 7 |
| 151 | 68 | 29.8 | 1.69 | 50 |

1-Body mass index in Kg/m<sup>2</sup> (BMI), Estimated body surface area (BSA) in m<sup>2</sup>. Colors: White – mild cases; Yellow – moderate cases; Orange – severe case.

**Figure 3A** shows a bee inside the ocular conjunctiva of participant, proving how aggressive this accident can be. Figure 3B shows a Africanized honeybee.

**Figure 3.**
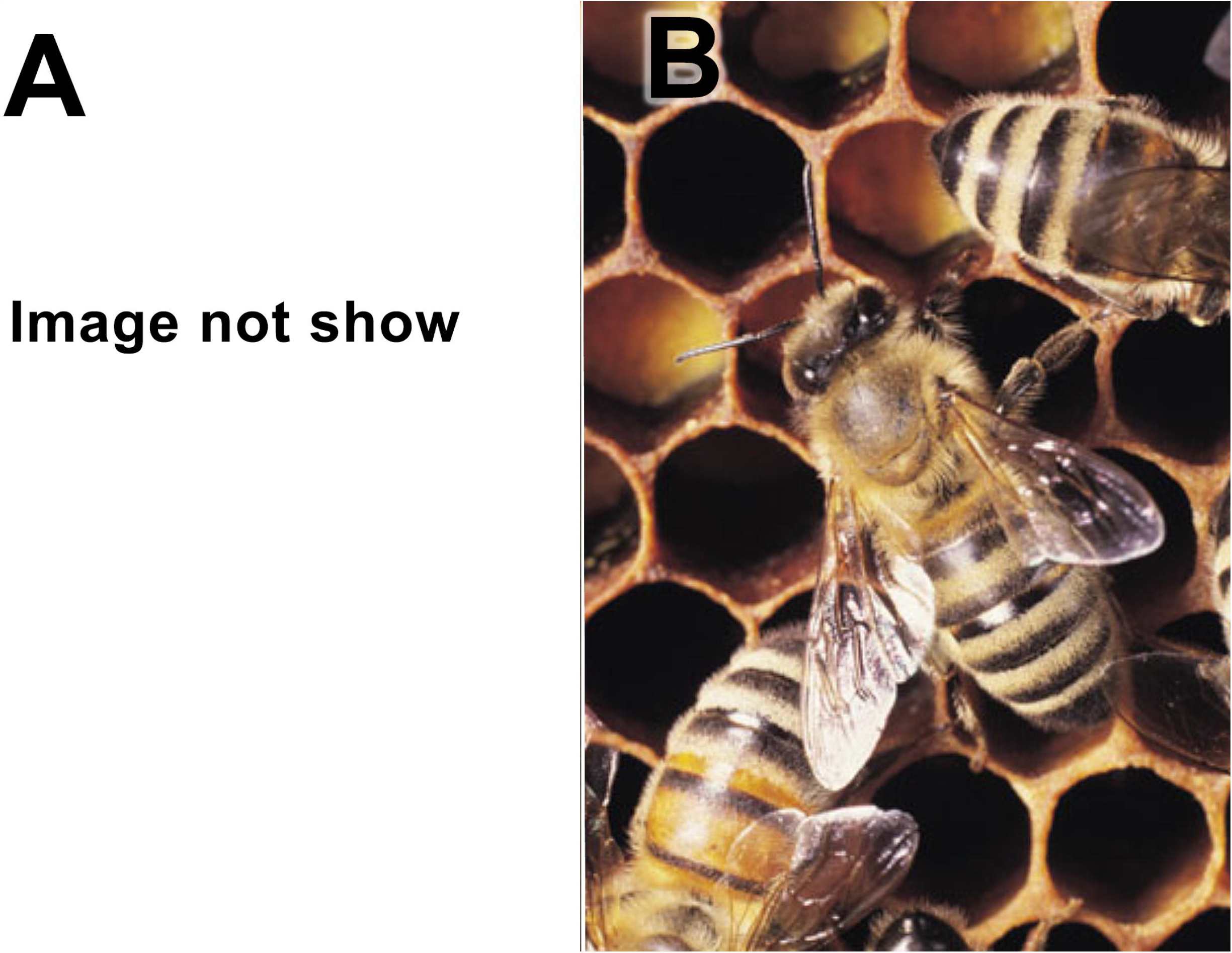
**(A)** Participant with *Apis mellifera* Africanized honeybee inside the ocular conjunctiva (Image not shown). **(B)** *Apis mellifera* Africanized honeybee.

### 3.2 Clinical outcome

#### 3.2.1 Adverse events not related to the investigational product

Participant 103 showed an abscess on the lower left flank (hypogastric region); 105 had inflammation of the right eye; 115 presented urine with strong smell, testicular and lumbar region (kidney) pain; 113 had tachycardia and edema in the lower limbs (chronic hypertensive patient), and 111 had bronchospasm. At follow-up visits, especially 10 days after treatment, all four participants, ranging from moderate to severe, complained of intense itching in the lesions. This complaint was also mentioned by one of the participants who suffered accident considered mild.

#### 3.2.2 Adverse events related to the investigational product

During the AAV infusion participant 110 presented numb lips and itchy head and 117 had pruritus and urticarial reaction. All with AEs related to the product were adequately treated and then the infusion of AAV was completed. All adverse events, either related or unrelated to the study, are described in **Additional File 3**.

### 3.3 Clinical and laboratory outcome

**Table 5** suggests that, from a clinical perspective, only one patient had hemodynamic alterations, whereas two presented respiratory disorders. From a laboratory perspective, 15 had high CPK levels, 9 showed increased levels of CRP, 8 had leukocytosis, 7 showed an increased fibrinogen levels and 4 in ALT. All results of laboratory tests for CPK, CRP, ALT, and complete blood count – including platelets, fibrinogen and creatinine – are available in the supplemental material in **Additional Files 4 and 5**.

**Table 5.**
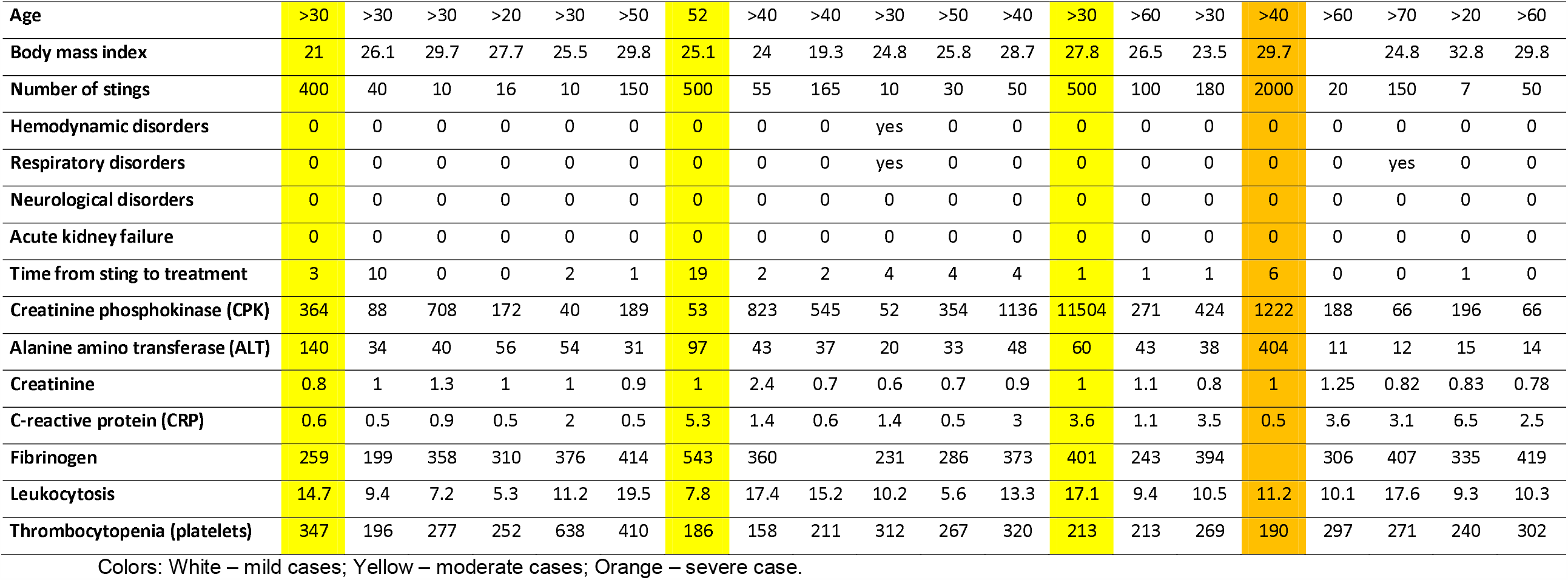
Distribution of clinical and laboratory alterations before treatment with Apilic Antivenom.

| Age | >30 | >30 | >30 | >20 | >30 | >50 | 52 | >40 | >40 | >30 | >50 | >40 | >30 | >60 | >30 | >40 | >60 | >70 | >20 | >60 |
| --- | --- | --- | --- | --- | --- | --- | --- | --- | --- | --- | --- | --- | --- | --- | --- | --- | --- | --- | --- | --- |
| Body mass index | 21 | 26.1 | 29.7 | 27.7 | 25.5 | 29.8 | 25.1 | 24 | 19.3 | 24.8 | 25.8 | 28.7 | 27.8 | 26.5 | 23.5 | 29.7 |  | 24.8 | 32.8 | 29.8 |
| Number of stings | 400 | 40 | 10 | 16 | 10 | 150 | 500 | 55 | 165 | 10 | 30 | 50 | 500 | 100 | 180 | 2000 | 20 | 150 | 7 | 50 |
| Hemodynamic disorders | 0 | 0 | 0 | 0 | 0 | 0 | 0 | 0 | 0 | yes | 0 | 0 | 0 | 0 | 0 | 0 | 0 | 0 | 0 | 0 |
| Respiratory disorders | 0 | 0 | 0 | 0 | 0 | 0 | 0 | 0 | 0 | yes | 0 | 0 | 0 | 0 | 0 | 0 | 0 | yes | 0 | 0 |
| Neurological disorders | 0 | 0 | 0 | 0 | 0 | 0 | 0 | 0 | 0 | 0 | 0 | 0 | 0 | 0 | 0 | 0 | 0 | 0 | 0 | 0 |
| Acute kidney failure | 0 | 0 | 0 | 0 | 0 | 0 | 0 | 0 | 0 | 0 | 0 | 0 | 0 | 0 | 0 | 0 | 0 | 0 | 0 | 0 |
| Time from sting to treatment | 3 | 10 | 0 | 0 | 2 | 1 | 19 | 2 | 2 | 4 | 4 | 4 | 1 | 1 | 1 | 6 | 0 | 0 | 1 | 0 |
| Creatinine phosphokinase (CPK) | 364 | 88 | 708 | 172 | 40 | 189 | 53 | 823 | 545 | 52 | 354 | 1136 | 11504 | 271 | 424 | 1222 | 188 | 66 | 196 | 66 |
| Alanine amino transferase (ALT) | 140 | 34 | 40 | 56 | 54 | 31 | 97 | 43 | 37 | 20 | 33 | 48 | 60 | 43 | 38 | 404 | 11 | 12 | 15 | 14 |
| Creatinine | 0.8 | 1 | 1.3 | 1 | 1 | 0.9 | 1 | 2.4 | 0.7 | 0.6 | 0.7 | 0.9 | 1 | 1.1 | 0.8 | 1 | 1.25 | 0.82 | 0.83 | 0.78 |
| C-reactive protein (CRP) | 0.6 | 0.5 | 0.9 | 0.5 | 2 | 0.5 | 5.3 | 1.4 | 0.6 | 1.4 | 0.5 | 3 | 3.6 | 1.1 | 3.5 | 0.5 | 3.6 | 3.1 | 6.5 | 2.5 |
| Fibrinogen | 259 | 199 | 358 | 310 | 376 | 414 | 543 | 360 |  | 231 | 286 | 373 | 401 | 243 | 394 |  | 306 | 407 | 335 | 419 |
| Leukocytosis | 14.7 | 9.4 | 7.2 | 5.3 | 11.2 | 19.5 | 7.8 | 17.4 | 15.2 | 10.2 | 5.6 | 13.3 | 17.1 | 9.4 | 10.5 | 11.2 | 10.1 | 17.6 | 9.3 | 10.3 |
| Thrombocytopenia (platelets) | 347 | 196 | 277 | 252 | 638 | 410 | 186 | 158 | 211 | 312 | 267 | 320 | 213 | 213 | 269 | 190 | 297 | 271 | 240 | 302 |

Colors: White – mild cases; Yellow – moderate cases; Orange – severe case.

The AHB score assigned to the participants before AAV administration showed level 7 for two, level 6 for five, and 5 or below 5 for 13 (**Table 6**). The most severe participant had a score of 7 and the three considered to be of moderate severity had respective scores of 7 in one and 6 in two. All 16 participants considered to be of mild severity had a score of 5 or below.

**Table 6.**
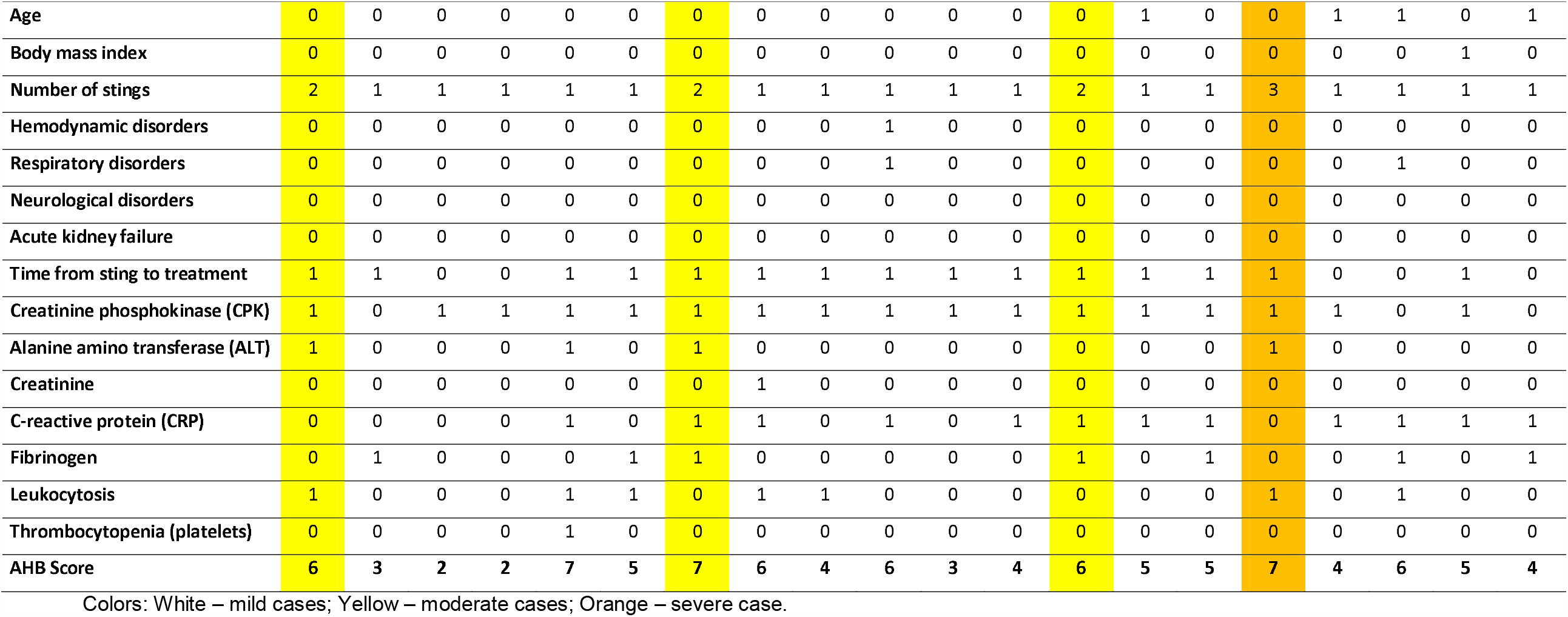
AHB Score before treatment, including the clinical parameters.

|  |  |  |  |  |  |  |  |  |  |  |  |  |  |  |  |  |  |  |  |  |
| --- | --- | --- | --- | --- | --- | --- | --- | --- | --- | --- | --- | --- | --- | --- | --- | --- | --- | --- | --- | --- |
| Age | 0 | 0 | 0 | 0 | 0 | 0 | 0 | 0 | 0 | 0 | 0 | 0 | 0 | 1 | 0 | 0 | 1 | 1 | 0 | 1 |
| Body mass index | 0 | 0 | 0 | 0 | 0 | 0 | 0 | 0 | 0 | 0 | 0 | 0 | 0 | 0 | 0 | 0 | 0 | 0 | 1 | 0 |
| Number of stings | 2 | 1 | 1 | 1 | 1 | 1 | 2 | 1 | 1 | 1 | 1 | 1 | 2 | 1 | 1 | 3 | 1 | 1 | 1 | 1 |
| Hemodynamic disorders | 0 | 0 | 0 | 0 | 0 | 0 | 0 | 0 | 0 | 1 | 0 | 0 | 0 | 0 | 0 | 0 | 0 | 0 | 0 | 0 |
| Respiratory disorders | 0 | 0 | 0 | 0 | 0 | 0 | 0 | 0 | 0 | 1 | 0 | 0 | 0 | 0 | 0 | 0 | 0 | 1 | 0 | 0 |
| Neurological disorders | 0 | 0 | 0 | 0 | 0 | 0 | 0 | 0 | 0 | 0 | 0 | 0 | 0 | 0 | 0 | 0 | 0 | 0 | 0 | 0 |
| Acute kidney failure | 0 | 0 | 0 | 0 | 0 | 0 | 0 | 0 | 0 | 0 | 0 | 0 | 0 | 0 | 0 | 0 | 0 | 0 | 0 | 0 |
| Time from sting to treatment | 1 | 1 | 0 | 0 | 1 | 1 | 1 | 1 | 1 | 1 | 1 | 1 | 1 | 1 | 1 | 1 | 0 | 0 | 1 | 0 |
| Creatinine phosphokinase (CPK) | 1 | 0 | 1 | 1 | 1 | 1 | 1 | 1 | 1 | 1 | 1 | 1 | 1 | 1 | 1 | 1 | 1 | 0 | 1 | 0 |
| Alanine amino transferase (ALT) | 1 | 0 | 0 | 0 | 1 | 0 | 1 | 0 | 0 | 0 | 0 | 0 | 0 | 0 | 0 | 1 | 0 | 0 | 0 | 0 |
| Creatinine | 0 | 0 | 0 | 0 | 0 | 0 | 0 | 1 | 0 | 0 | 0 | 0 | 0 | 0 | 0 | 0 | 0 | 0 | 0 | 0 |
| C-reactive protein (CRP) | 0 | 0 | 0 | 0 | 1 | 0 | 1 | 1 | 0 | 1 | 0 | 1 | 1 | 1 | 1 | 0 | 1 | 1 | 1 | 1 |
| Fibrinogen | 0 | 1 | 0 | 0 | 0 | 1 | 1 | 0 | 0 | 0 | 0 | 0 | 1 | 0 | 1 | 0 | 0 | 1 | 0 | 1 |
| Leukocytosis | 1 | 0 | 0 | 0 | 1 | 1 | 0 | 1 | 1 | 0 | 0 | 0 | 0 | 0 | 0 | 1 | 0 | 1 | 0 | 0 |
| Thrombocytopenia (platelets) | 0 | 0 | 0 | 0 | 1 | 0 | 0 | 0 | 0 | 0 | 0 | 0 | 0 | 0 | 0 | 0 | 0 | 0 | 0 | 0 |
| AHB Score | 6 | 3 | 2 | 2 | 7 | 5 | 7 | 6 | 4 | 6 | 3 | 4 | 6 | 5 | 5 | 7 | 4 | 6 | 5 | 4 |

Colors: White – mild cases; Yellow – moderate cases; Orange – severe case.

The laboratory tests showed an increase in CPK in 15 participants, CRP in 9, leukocytosis in 8, fibrinogen in 7 and ALT in 4. The biological AHB score, excluding clinical parameters, assigned to participants before AAV administration, showed level 4 in three participants, level 3 in five and level 2 or lower in 13 (**Table 7**).

**Table 7.** AHB Score before treatment, excluding the clinical parameters.

|  |  |  |  |  |  |  |  |  |  |  |  |  |  |  |  |  |  |  |  |  |
| --- | --- | --- | --- | --- | --- | --- | --- | --- | --- | --- | --- | --- | --- | --- | --- | --- | --- | --- | --- | --- |
| **Creatinine phosphokinase (CPK)** | 364 | 88 | 708 | 172 | 40 | 189 | 53 | 823 | 545 | 52 | 354 | 1136 | 11504 | 271 | 424 | 1222 | 188 | 66 | 196 | 66 |
| **Alanine amino transferase (ALT)** | 140 | 34 | 40 | 56 | 54 | 31 | 97 | 43 | 37 | 20 | 33 | 48 | 60 | 43 | 38 | 404 | 11 | 12 | 15 | 14 |
| **Creatinine** | 0.8 | 1 | 1.3 | 1 | 1 | 0.9 | 1 | 2.4 | 0.7 | 0.6 | 0.7 | 0.9 | 1 | 1.1 | 0.8 | 1 | 1.25 | 0.82 | 0.83 | 0.78 |
| **C-reactive protein (CRP)** | 0.6 | 0.5 | 0.9 | 0.5 | 2 | 0.5 | 5.3 | 1.4 | 0.6 | 1.4 | 0.5 | 3 | 3.6 | 1.1 | 3.5 | 0.5 | 3.6 | 3.1 | 6.5 | 2.5 |
| **Fibrinogen** | 259 | 199 | 358 | 310 | 376 | 414 | 543 | 360 |  | 231 | 286 | 373 | 401 | 243 | 394 |  | 306 | 407 | 335 | 419 |
| **Leukocytosis** | 14.7 | 9.4 | 7.2 | 5.3 | 11.2 | 19.5 | 7.8 | 17.4 | 15.2 | 10.2 | 5.6 | 13.3 | 17.1 | 9.4 | 10.5 | 11.2 | 10.1 | 17.6 | 9.3 | 10.3 |
| **Thrombocytopenia (platelets)** | 347 | 196 | 277 | 252 | 638 | 410 | 186 | 158 | 211 | 312 | 267 | 320 | 213 | 213 | 269 | 190 | 297 | 271 | 240 | 302 |
| **AHB Score** | 3 | 1 | 1 | 1 | 5 | 3 | 4 | 4 | 2 | 2 | 1 | 2 | 3 | 2 | 3 | 3 | 2 | 3 | 2 | 2 |

Colors: White – mild cases; Yellow – moderate cases; Orange – severe case.

**Table 8** reveals that none of the participants showed clinical alterations 30 days after administration of AAV. However, the following laboratory tests were changed: CRP in four, fibrinogen in three, CPK in two, ALT and leukocytosis in one.

**Table 8.**
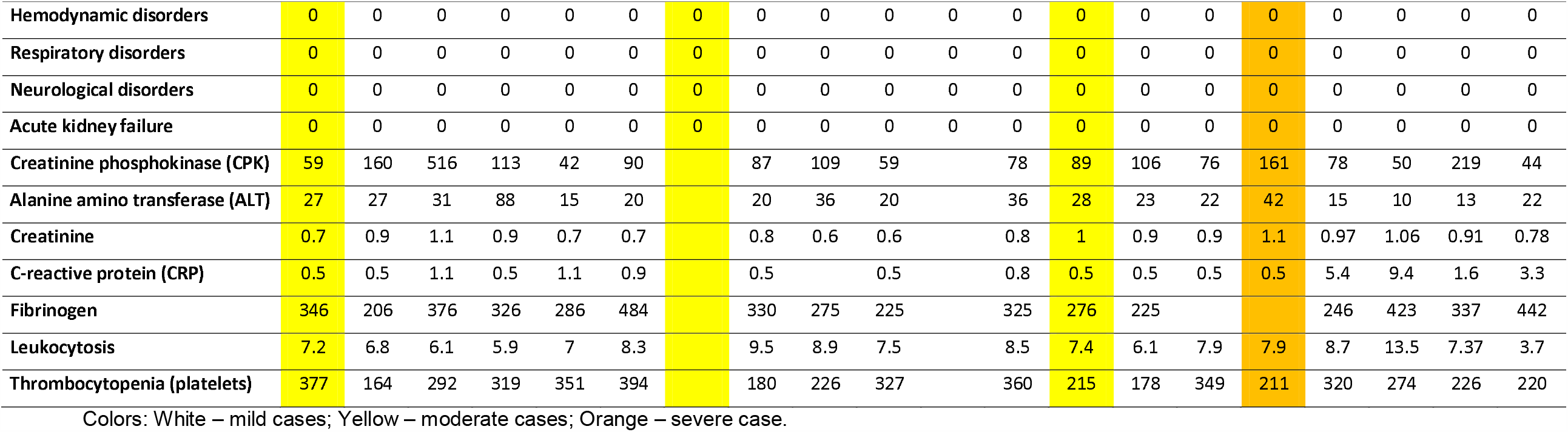
Distribution of clinical and laboratory alterations 30 days after treatment.

**Table 9** shows eight participants still had changes in laboratory AHB score 30 days after AAV administration, distributed as follow: one at level 3, one at level 2 and six at level 1. Four participants, including three considered moderate and one severe, presented a laboratory AHB score of zero.

**Table 9.**
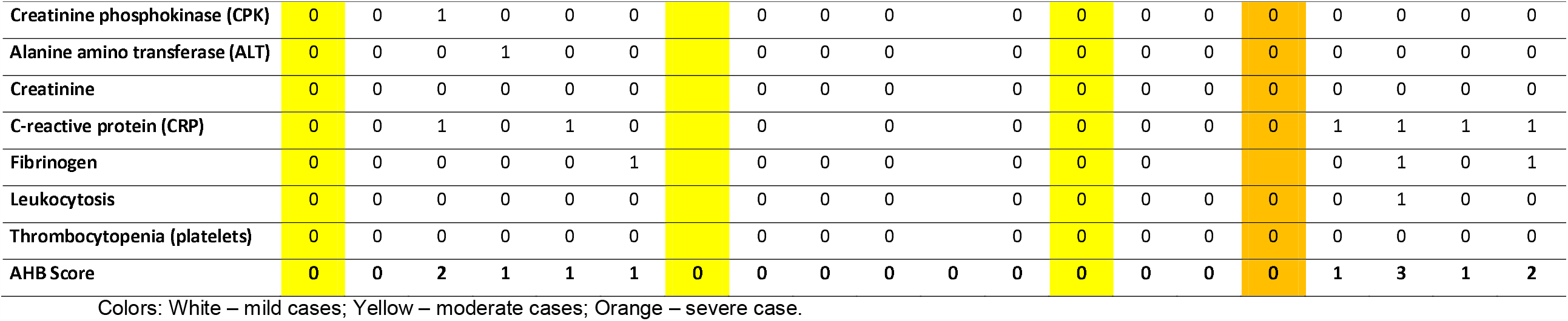
AHB score 30 days after treatment, excluding the clinical parameters.

### 3.4. Melittin and phospholipase A_2_ pharmacokinetics - ELISA assays

All participants were monitored at times zero, two, six, 12, 24- and 48-hour after inclusion during hospitalization, and 10, 20 and 30 days during external follow-up. Of the 20 participants, 6 participants had missed at least one of the scheduled sample collection schedules, and two participants (301 and 303) did not perform the ELISA test.

Fourteen participants had a complete follow-up and all blood samples were obtained. Concentrations of melittin + phospholipase A_2_ varied between 0.03 ng/mL and 587.35 ng/mL during hospitalization and during the follow-up surveillance, despite the excellent clinical conditions of all the participants, melittin + phospholipase A_2_ concentrations ranged between 0 and 1.479 ng/mL.

The participant 101 suffered approximately 400 stings, and when attended he received 6 vials of AAV three days after the accident (**Figure 4A**). At ten, twenty and thirty days after AAV administration, during outpatient follow-up, the respective levels of melittin + phospholipase A_2_ were 709, 497 and 273 ng/mL.

**Figure 4.**
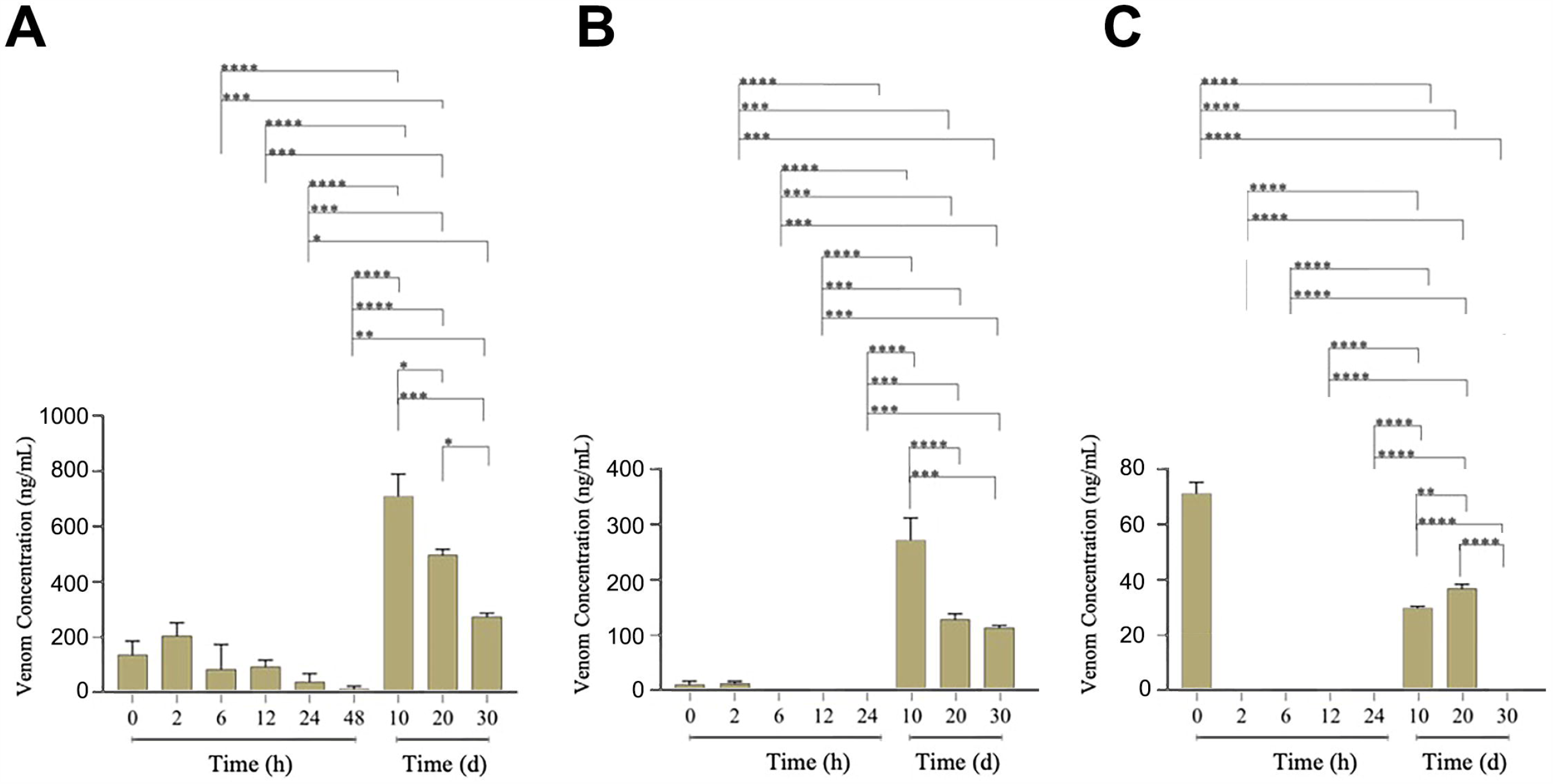
Quantification of *Apis mellifera* venom in the blood of Participant **(A), (B)** and, **(C)** using ELISA test. Time 0 corresponds to the patient’s admission to the hospital. Times 2, 6, 12, 24 and 48 h refer to the time elapsed after AAV administration. The times 10, 20, and 30 days correspond to the patient follow-up after discharge from hospital. The values represent the mean ± standard deviation of the absorbances of each biological replica. The results obtained were compared using the ANOVA test for repeated measures, followed by Tukey test (*p<0.1; **p<0.01; ***p<0.001 and ****p<0.0001).

The participant 103 suffered about 10 stings, and when attended he received two vials of AAV on the first day after the accident (**Figure 4B**). Ten, twenty and thirty days after, at outpatient return visits, the respective melittin + phospholipase A_2_ levels were 276, 126 and 114 ng/L.

The participant 302 suffered approximately 150 stings, and when attended he received two vials of AAV on the first day after the accident (**Figure 4C**). Ten, twenty and thirty days after, at outpatient return visits, the respective melittin + phospholipase A_2_ levels were 29, 36, and zero ng/mL.

**Figure 4** (A, B, C) shows the course of melittin + phospholipase A_2_ levels in participants 101, 103 and, 302 at hospital admission and, two, six, 12, 24- and 48-hour after inclusion during hospitalization, and 10, 20 and 30 days during external follow-up

All the results of ELISA assays are available in **Additional File 6**.

### 3.5 Mass spectrometry

Analyses of the TIC chromatograms (**Figure 5A, 5B**), indicating the 712.15 ion [M+4H^4+^] as well as the MS^2^ profiles, particularly the [y_13_]^2+^ fragment made it possible to determine the presence and relative level of melittin in participants, according to **Table 10**. The determination of the relative melittin quantity in the serum was performed according to the graphical interpretation of the spectra, as presented in supplemental material at **Additional File 2**.

**Table 10.** Qualitative melittin detection in 19 participants 30 days after the AAV treatment.

|  |  |  |  |  |  |  |  |  |  |  |
| --- | --- | --- | --- | --- | --- | --- | --- | --- | --- | --- |
| **Number of stings** | 400 | 40 | 10 | 16 | 10 | 150 | 500 | 55 | 165 | 10 |
| **Number of vials** | 6 | 2 | 2 | 2 | 2 | 2 | 6 | 2 | 2 | 2 |
| **Presence of melittin** | (+) | - | - | - | (+) | - | (*) | (+) | - | (++) |
| **Number of stings** | 30 | 50 | 500 | 100 | 180 | 2.000 | 20 | 150 | 7 | 50 |
| **Number of vials** | 2 | 2 | 6 | 2 | 2 | 10 | 2 | 2 | 2 | 2 |
| **Presence of melittin** | - | - | (+) | (+++) | - | (+) | - | - | - | (++) |

\* (Not evaluated). Colors: White – mild cases; Yellow – moderate cases; Orange – severe case.

**Figure 5.**
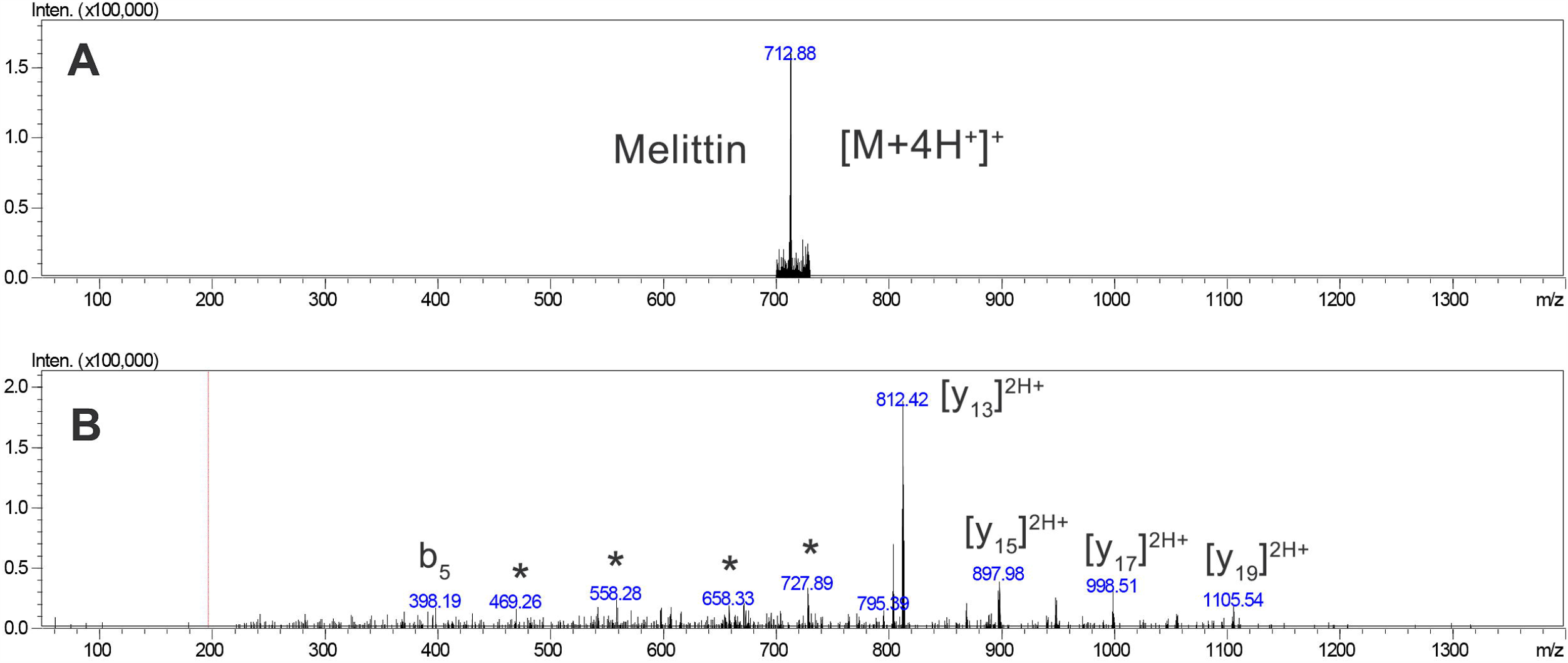
Representative mass spectrometry profile of the qualitative assessment of the presence of melittin in the serum of participant 115 considered positive (+++): **(A)** Melittin [M+4H]4+ MS profile and (B) MS^2^ interpreted profile, annotated for the larger b and y ions, as well as internal fragments (*).

## 4. Discussion

According to Bochner (36), *the discovery of antivenom serotherapy, which occurred in France, was presented to the French Society of Biology on February 10, 1894, by representatives of two Parisian research institutions. The organizations were the National Museum of Natural History, represented by Césaire Auguste Phisalix and Gabriel Bertrand, under the direction of Jean Baptiste Auguste Chauveau, and the Pasteur Institute, represented by Albert Calmette, under the direction of Emile Roux* (37, 38). *In Brazil, the discovery of antivenom serotherapy had a profound impact on the work of Vital Brazil Mineiro da Campanha (1865–1950), a researcher known worldwide for his scientific discoveries and for his evidence of the specificity of antivenom serums. He was also responsible for creating the Butantan Institute (BI) in São Paulo city, São Paulo state, and the Vital Brazil Institute (VBI) in Niteroi, Rio de Janeiro state* (39-41).

Since the 2000s CEVAP started the development of new antivenoms including a new Apilic Antivenom (AAV) denominated by Pucca *et al*. (5) as “*a next-generation antivenom*”. This new approach is based on its development just precisely against the two main toxins from the Africanized (*Apis mellifera*) honey bee venom, namely, melittin and phospholipase A_2_ (21). Allergens and nociceptive components were removed from the crude venom, which significantly reduced the suffering of the serum-producer animals, thus allowing the immune response of the herd to be directed towards what is really toxic and harmful to the envenomed human being. Then, after the validation of the candidate, researchers from two Brazilian antivenom producers - *Vital Brazil Institute (VBI)* and *Butantan Institute (BI) -* joined the team to produce a new AAV for clinical and pre-clinical trials.

In 2017, the Brazilian National Health Surveillance Agency (ANVISA), through Resolution RDC N ° 187 of November 8, 2017, established the minimum requirements for the registration of hyperimmune sera, aiming to guarantee the quality, safety, and efficacy of these products. It also established that the registration petition of these immunobiologicals must include the results of clinical trial reports. This is because until then all Brazilian antivenoms had not been previously validated by clinical trials (42). Therefore, this I/II clinical trial using the AAV for treating of massive Africanized honeybee (*Apis mellifera*) attacks, whose clinical protocol was published in 2017 by Barbosa et al. (23), is the first following this new guideline.

According to the WHO Guideline (43) clinical trials with antivenoms are designed to address three main issues: (a) the optimal initial dose; (b) efficacy, i.e. the ability of antivenom to control the main clinical manifestations of envenoming; and (c) safety, i.e. the incidence and severity of early and late adverse reactions. Dose-finding studies are usually followed by randomized, controlled trials in which the new antivenom is compared to another antivenom already used or, in its absence, two doses of the new antivenom are compared (44).

As AAV was deliberately targeted to neutralizes the two major and main toxic compounds of venom, the clinical effect of low-mass proteins and peptides that cause, for example, intense pruritus, flushing, hyperthermia, papules, urticarial plaques, hypotension, tachycardia, headache, nausea, vomiting, abdominal colic, bronchospasms, and psychomotor agitation, were neutralized by symptomatic medications such as antihistamines, corticosteroids, adrenaline (when the anaphylactic shock is suspected), and pethidine hydrochloride for severe pain. In the presence of bronchospasm, oxygen (O_2_) catheter-associated with inhaled bronchodilators of the type β-2- agonists (salbutamol, fenoterol, or terbutaline) was used in the usual recommended doses (23). As the main outcome of the study was to assess safety, including the severity of acute adverse events, and to confirm the lowest effective dose when confronted with different inoculated amounts of venom, the main adverse events caused by equine-heterologous serotherapy were studied.

Safety was assessed mainly by early adverse events, particularly IgE-mediated anaphylactic reactions type I, non-IgE-mediated anaphylactic reactions, pyrogenic reactions (endotoxin contamination) and late adverse reactions (type III hypersensitivity, such as serum sickness) (45-48).

Among the adverse events unrelated to the product, the persistence of itching at the lesions of sting site at the first follow-up visit, i.e. 10 days after AAV administration, should be highlighted. This clinical picture and its persistence at the sting site or in the subcutaneous cellular tissue is related to the envenoming (4, 6, 7, 18). Since the clinical protocol recommended the use of antihistamines only during the hospitalization period, this point should be revised and the use of this medication should be extended for at least 15 days.

Regarding the adverse events related to the product, only two participants (10%) experienced mild and early adverse reactions such as numbness of the lips, itchy head, pruritus, and urticarial reaction. Recently, Mendonça-da-Silva et al. (49) evaluated the safety and efficacy of a freeze-dried trivalent antivenom for snakebites in the Brazilian Amazon, where 112 participants were treated in an open-label, randomized controlled phase IIb clinical trial. Twenty-three (19.8%) participants showed early adverse events after antivenom therapy. The most common were urticaria (13.8%), pruritus (11.2%), facial flushing (3.4%), and vomiting (3.4%). Our results agreed with those of Mendonça-da-Silva et al. (49) and with other studies in the world literature (45-48). None of the participants presented with serum sickness. AAV appears to be a safe investigational product for clinical use.

It should be emphasized that in the event of an accident with AHB, adverse reactions classically described for the antivenoms might be confused with adverse events of envenoming itself. Therefore, it becomes hard to define which was the actual cause, whether it was the venom or the AAV (4, 6, 7, 18). Despite these limitations, it was possible to conclude that the adverse events related to the investigational product were similar to those due to common antivenoms used for other types of envenoming (45-49), thus confirming the safety of AAV for clinical use.

In relation to the clinical and laboratory outcome, the authors proposed the new AHB Score to assess the severity of the participants at the moment of the first visit, in addition to the evolutive outcome. The use of the AHB score proved to be simple and showed that it is proportional to the severity of the envenomation and to the normalization of clinical and biological signs. Combining clinical and biological criteria, the score appears both sensitive and accurate, in particular the biological variables, to characterize the severity and course of envenomation. Thus, this allowed us to confirm clinical cure in all participants 30 days after AAV administration. However, biological disturbances remained in some participants (8/20) who, however, did not have particularly severe envenomation. Surprisingly, the two most envenomed participants who received 500 and 2000 injections and who were assigned an AHB score of 7 had a score of zero 30 days after AAV administration. In addition, out of the 5 participants who had an AHB score of 6 at inclusion, all had a score of zero on thirty days but one remained at 1 because of a CRP level slightly above the limit (1.1 mg/dl). It should also be emphasized that envenoming’s caused by venomous animals trigger the systemic inflammatory response syndrome described starting in the 1990s by several authors (50-54). This constitutes an acute phase reaction with a massive release of pro-inflammatory cytokines, particularly IL-1, IL-6, and TNF alpha, and acute-phase proteins, especially C-reactive protein. These acute-phase reactions observed are in accordance with previous clinical studies with other antivenoms (4, 6, 50-55).

The ELISA test, standardized to assess the presence of melittin and phospholipase A_2_ during the study, showed the reappearance of these two fractions 10, 20, and 30 days after treatment in most of participants.

When studying an ovine model to evaluate the interactions between the venom of *Micrurus fulvius* and F(ab’)_2_ antivenom administered intravenously, Paniagua et al. (56) observed immediate neutralization of the venom in the bloodstream. Antivenom also appears not to be available to neutralize the residual venom deposited at the site of injection, so the venom can remain active with slow transfer to the bloodstream for ongoing systemic distribution (56). However, as long as the antivenom remains available in the plasma compartment, the venom is captured and eliminated until the plasma concentration of antivenom is no longer sufficient to neutralize the released venom (57-59). This phenomenon is well illustrated by the use of Fab - which has a short half-life of around 10-15 hours (57) - in the treatment of envenomation by Crotalidae which requires the recurrent renewal of the administration of Fab antivenom (60, 61). Under these conditions, the renewal of the antivenom serum must be planned according to, on the one hand, the half-life of the F(ab’)_2_ antibodies, approximately 50 hours (57), and on the other hand, the quantity of bee venom in circulation, which is more difficult to determine.

We hypothesis that, in agreement with Paniagua et al. (56), the multiplicity of bee sting sites occur in many parts of the human body depositing the venom in the subcutaneous tissue from where it is slowly released into the blood. In addition to the clearance of the F(ab’)_2_ molecule, the latter remains in the plasma compartment without entering the tissues, preventing the neutralization of the venom in the subcutaneous cellular tissue (58). Surprisingly, the participants that received only two vials of AAV were those who still presented a systemic inflammatory response syndrome 30 days after treatment, i.e. an inflammatory response based on augmented levels of CRP (4 participants), fibrinogen (3 participants), and leukocytes in one (4, 50-55). This can be explained by a lower concentration of F(ab’)_2_ which is eliminated more quickly enabling the venom, even in small quantities, to stay longer in the body. This clearly raises the question of renewing AAV in participants who suffered multiple AHB stings a week after the first administration or increasing the number of vials at the time of the first attendance, even if the initial symptoms are mild.

Finally, qualitative mass spectrometry (8) showed the presence of melittin in the blood of mild (five), moderate (two), and severe (one) participat 30 days after AAV administration. It should be emphasized that participant 115, who received two vials of AAV for being stung by approximately 100 honeybees, experienced a particular increase in circulating melittin 30 days after treatment. Thus, the clinical response of the participants, laboratory tests from the acute-phase response, ELISA test, and finally mass spectrometry, allows us to consider that although the investigational product is safe, i.e. the main objective of this study, it would be necessary to revise the clinical protocol, especially the number of vials of AAV to use. A multicenter phase III clinical trial should be performed to confirm these hypotheses and adjust the doses of this new antivenom.

## 5. Conclusions

The AAV proved to be safe, as related adverse events were observed in only two (10%) participants, corroborating reports of heterologous antivenoms use. No late adverse events were observed on days 10, 20, or 30 of the clinical surveillance. Preliminary efficacy was observed by clinical improvement in participants, decrease in acute-phase-reaction markers, and reduction in circulating melittin levels, and phospholipase A_2_ measured by ELISA. The doses recommended in the clinical protocol should be reassessed and increased, given that melittin was found, through mass spectrometry, in the blood of eight participants 30 days after the specific treatment. It should be noted that these issues were expected because it is a particular accident, different from all others described in the world caused by venomous animals. A phase III clinical trial should be performed to confirm these observations, adjust the recommended doses, and assess the product’s efficacy.

## Data Availability

The additional files will be able just on Article Published!

## Author Contributions

ANB, FST, and MBM were the principal investigators; FCTC, BCM, and LDS standardized the ELISA assay; RSF Jr and LERC produced and quality controlled the Apilic Antivenom; JNB, DJT, NBM, CVC, MTRC and APP recruited the participants; DCP developed the mass spectrometry analyses; LB, JPC, and BB discussed the proposal and corrected the manuscript. All authors have read and agreed to the published version of the manuscript.

## Funding

This study was supported by the National Council of Technological and Scientific Development (CNPq) under Grants No. 437089/2018-5 (LDS), 563582/2010-3 (BB), 401170/2013-6 (BB), and in part by grants from CAPES (Coordination for the Improvement of higher Education Personnel) [AUX-PE Toxinology Proc. No. 23,038.000823/201121]. RSF Jr. is a CNPq PQ1C fellow researcher [303224/2018-5]. DCP is a CNPq fellow researcher [301974/2019- 5].

### Acknowledgments

Special thanks to the Center for the Study of Venoms and Venomous Animals (CEVAP) from São Paulo State University (UNESP), Brazil. Pasqual Barreti, Rodrigo Bazan, Carlos Antonio Caramori and Patricia Carvalho Garcia from Botucatu Medical School from São Paulo State University (UNESP), Brazil. André Balbi and Emergency Room Team from the Botucatu Medical School Clinical Hospital (HCFMB), São Paulo, Brazil. Sergio Muller of the São Paulo State Department of Health. Camile Giaretta Sachetti, Patricia de Souza Boaventura and Felipe Nunes Bonifácio from the Department of Science and Technology of the Brazilian Ministry of Health. Ricardo de Oliveira Orsi from the Botucatu School of Veterinary Medicine and Zootechnics (UNESP), Brazil.

